# “Genomic and phenotypic characterization of Investigator Global Assessment (IGA) scale based endotypes in atopic dermatitis”

**DOI:** 10.1101/2020.04.16.20058677

**Authors:** Sandra P Smieszek, Bartlomiej Przychodzen, Sarah E Welsh, Jennifer Brzezynski, Alyssa Kaden, Michael Mohrman, Jingyuan Wang, Changfu Xiao, Sonja Ständer, Gunther Birznieks, Christos Polymeropoulos, Mihael H Polymeropoulos

## Abstract

**Background:** Atopic dermatitis (AD) is a heritable and heterogeneous inflammatory chronic skin disorder. Utilizing decision tree/supervised learning of extensive clinical, molecular and genetic data, we aimed to define distinct AD endotypes.

**Methods:** Deep phenotyping and whole-genome sequencing was performed on samples obtained from participants of EPIONE, a randomized-controlled phase III study in AD patients with severe pruritus comprising mild (23%), moderate (64%) and severe (13%) AD as determined by AD Investigator Global Assessment scale. Three categories of analysis were performed: clinical associations, lab value associations (EOS, IgE, cytokines) and genetic analysis of whole-genome sequencing data

**Results:** Based on a decision tree, we found that five clinical features from the SCORing Atopic Dermatitis (SCORAD) Index can accurately differentiate between IGA severities. We observe a significant difference between severity and eosinophil counts (p<0.001), IgE (p<0.001) and *Filaggrin (FLG*) LOF frequency (OR 2.3, CI 1.6-3.2, p<0.0001) as well as interleukin pathway genes, specifically IL5RA variants differentiating the groups.

**Conclusion:** Our results suggest significant differences between severity groups across a number of features appear to constitute distinct endotypes with likely distinct causative factors. Differing underlying pathophysiology’s indicate endotype knowledge is critical to help guide therapeutic approaches to AD.

**Capsule summary:**

- AD is a heritable and heterogeneous skin disorder that makes the ‘one size fits all’ therapeutic approach suboptimal for patients with AD.
- We attempted to define AD endotypes based on clinical, molecular, and genetic characteristics. Clinical, CBC, and genetic associations all tend to suggest existence of separate endotypes.

## Introduction

Atopic dermatitis (AD) is a complex and heterogeneous, highly heritable, chronic-relapsing, inflammatory skin disorder. AD affects 7.3% of US adults^1^,^2^. The genetic background of AD is complex and likely composed of a mixture of rare and common variants. The heritability of AD is estimated to be around 75%^2^. Studies have shown that there are genetic factors specific to AD beyond those for general atopy^3,4^. AD is manifested by a wide range of symptoms and significantly impacts quality of life. Flares are usually characterized by extreme pruritus of red, rough, flaky and often fissured regions of the skin, that over time thicken^2^. Chronic pruritus, lasting more than 6 weeks, is a key feature associated with AD. The extent of skin lesions and other physical symptoms vary within this patient population and can be diagnosed as mild, moderate, or severe as defined by the Investigator Global Assessment scale for Atopic Dermatitis (vIGA-AD™). The etiology is likely resultant from an interaction between the genetically determined skin barrier, immune system dysregulation, and the environmental triggers^2^.

One of the main functions of the skin is to act as a barrier between the individual and the environment, preventing water loss and at the same time preventing pathogen and allergen entry^5^. Skin barrier dysfunction is a key clinical feature of AD, as this facilitates penetration of allergens, immunological dysfunction, and consequently an increased risk of developing eczema^5^,^6^. The skin barrier dysfunction has, among others, been associated with the etiology of the itch-scratch cycle^7^. In addition, genes encoding skin barrier proteins have been shown to play a role in the heritability of AD^8^,^9^. Loss-of-function (LOF) variants resulting in aberrant *FLG* production, constitute the best-known AD gene-association and have been shown to predispose individuals to AD^10^,^11^. Individuals harboring *FLG* LOF variants manifest clinical features such as dry and fissured skin. LOF variants in *FLG* are associated with lower levels of natural moisturizing factors in AD^12^. Additional genes involved in skin barrier function, including epidermal differentiation complex (EDC) proteins, are thought to have a potential role in AD. The EDC encodes proteins critical to the proper development of keratinocytes and normal formation of the skin barrier^13^. Downregulation of EDC genes has also been implicated in AD^14,15,16,17^.

Disrupted skin barrier promotes penetration of allergens, immunological dysfunction, and consequently an increased risk of developing eczema^18^. Critical features of the TH2 immune response include local production of TH2 cytokines activation of eosinophils (EOS) and mast cells, as well as production of IgE^19^. Ultimately T-cell subsets, including Th2/Th22 cells, are upregulated in AD skin^20^. Interestingly both lesional and nonlesional AD showed a highly polyclonal T cell receptor pattern, insinuating the lasting presence of potentially pathogenic memory T cells beyond clinically inflamed lesions^20^. Ultimately dysregulation leads to increased production of neuropeptides such as substance P (SP) released from primary sensory nerves in the skin^21^. SP binds the neurokinin-1 receptor (NK1R) and MRGPRS^22^. Elevated SP levels have been observed in the blood and lesional skin of AD patients, and have been correlated with itch intensity, as shown in challenge studies and further reiterated in transcriptome analysis ^23^,^7^. Elevated SP levels further elicit vasodilation of short duration leading to mast cell degranulation, nerve growth factor expression in keratinocytes, and neurogenic inflammation associated with erythema and pruritus^21^. Proof of concept studies have shown increases in itch resulting from injected SP and expression of NK1R in keratinocytes, as well as reduction in itch resulting from inhibition of NK1R. Pruritus, perpetuates scratching that further damages the skin barrier^7^.

Our work aimed to evaluate the effect of clinical manifestations, molecular levels and genetic associations and cytokine levels to further understand the architecture and variability of AD as assessed in subjects during baseline visit. We aim to discern whether degree of severity recapitulates individual endotypes in AD.

## Results

EPIONE, a multi-center, placebo-controlled, double-blind study, enrolled and randomly assigned (1:1) 375 patients aged 18 to 70 with AD. Welsh et al., 2020 (manuscript under review) describes the efficacy and safety results from EPIONE. At baseline, participants presented with a range of disease severities from mild (23%) to moderate (64%), and severe (13%). Severity was defined by Investigator Global Assessment for Atopic Dermatitis (vIGA-AD™). For analysis, milder AD was defined as IGA scores of 1 (1) or 2 (77) at baseline, while more severe AD was defined as IGA scores of 3 (218) or 4 (33). In addition to deep phenotyping we obtained AD samples for whole genome sequencing (n=765 entire study population, n=332 randomized patients). **Table 1** shows demographic and clinical characteristics of the studied cohort at baseline. **Figure 1** depicts the baseline clinical and lab measurements across IGA groups. We examine each sub-component in depth in the sections that follow.

**Table 1.**
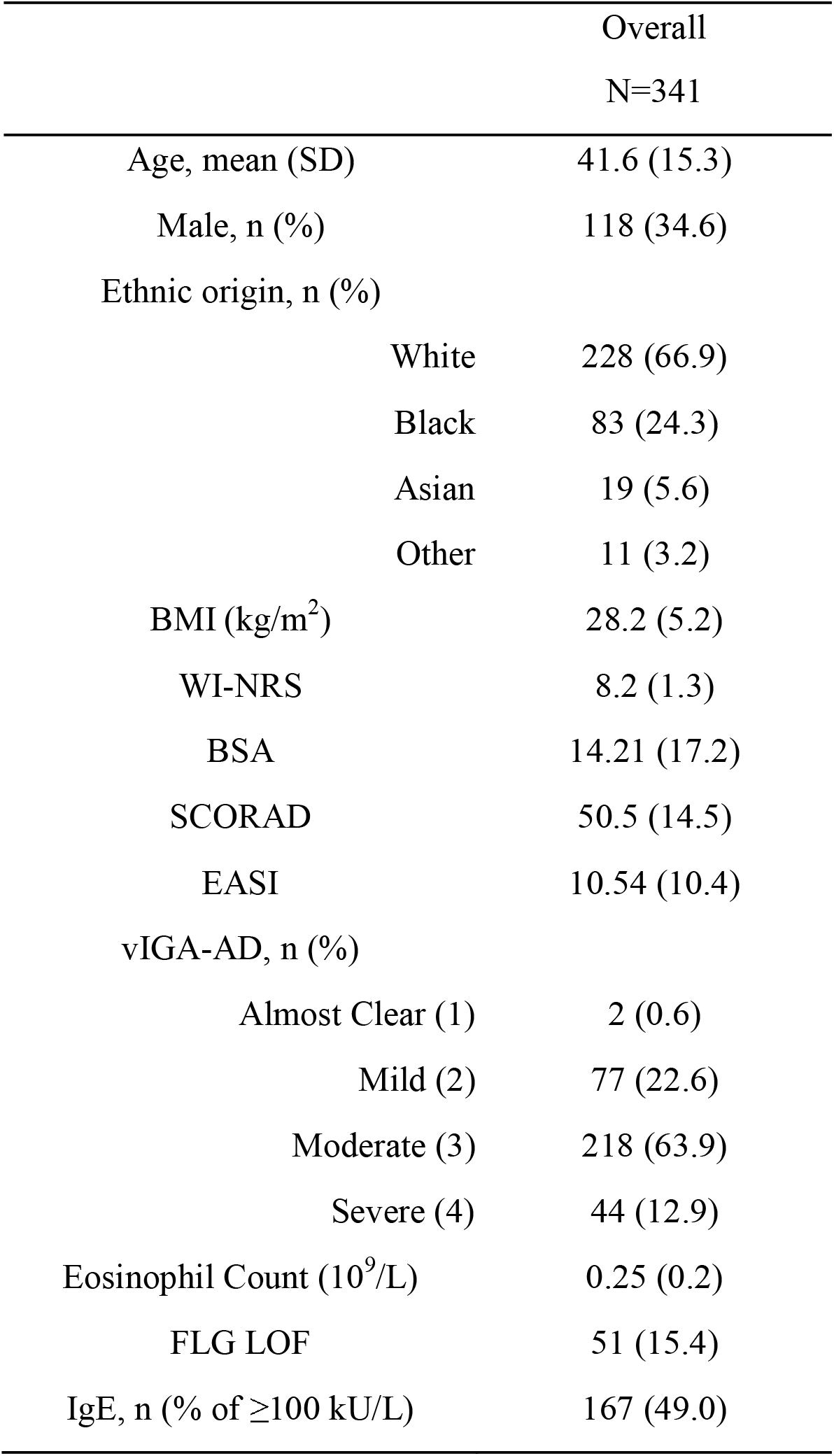
Baseline demographic and clinical characteristics.

**Figure 1.**
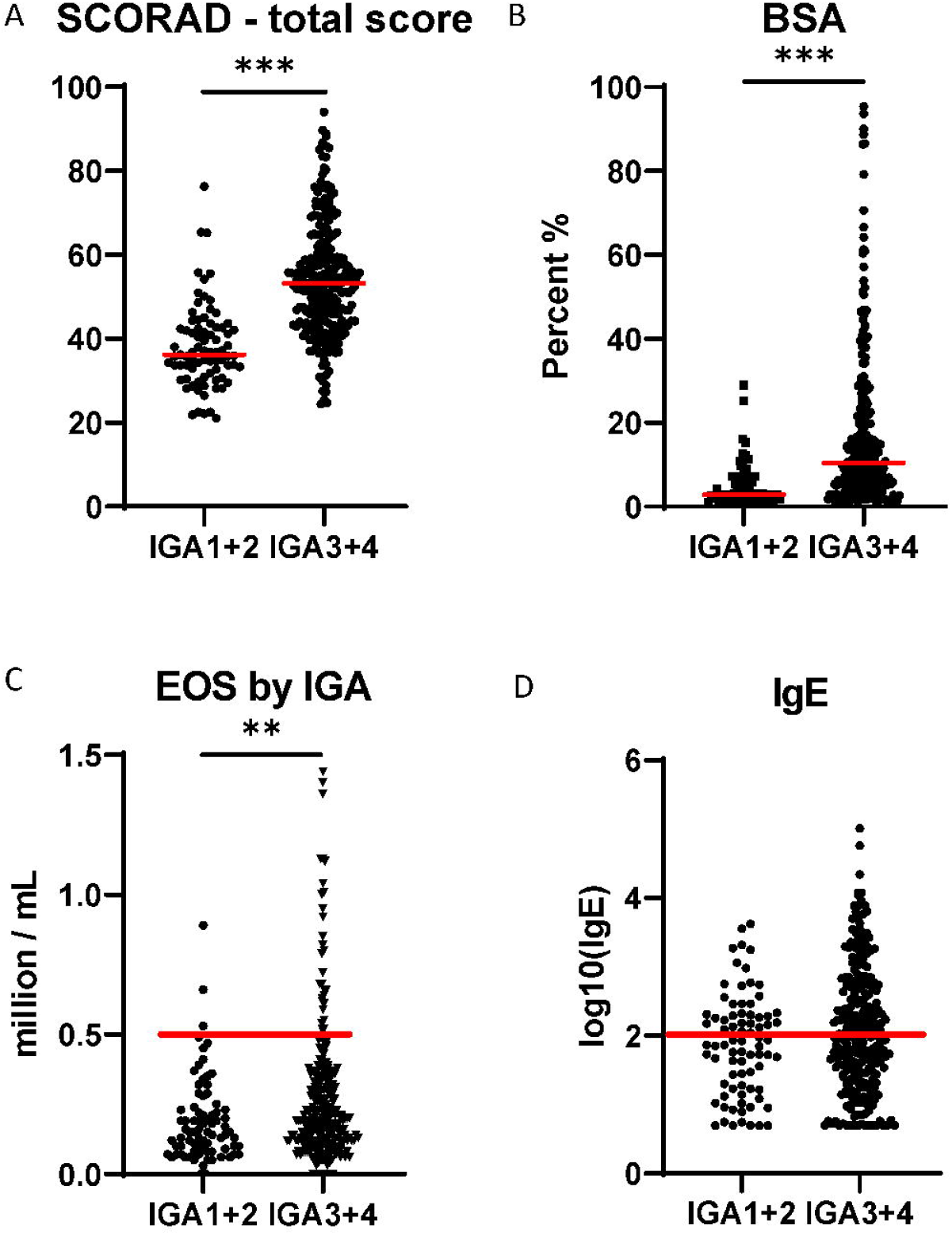
Atopic dermatitis endotypes.

### Clinical Association

We collected extensive clinical information using the SCORAD index including total area involved, intensity, and subjective measurements^24^,^25^. As expected, we observe that mild AD presents with fewer lesions, mild erythema, and minimal induration/papulation or oozing/crusting. In contrast, severe AD presents with a larger number of erythematous lesions associated with significant induration/papulation, oozing/crusting. Importantly, we observed that the interaction between three most informative variables (Edema/Papules, Erythema and Total Area Affected) can classify patients by disease severity. The most severe patients present almost exclusively with highest SCORAD Edema/Papules score (3 on a 0-3 scale) or SCORAD Erythema (3 on a 0-3 scale). Utilizing only those two variables we can accurately identify 35.8% of IGA3/4 with only 2.5% of IGA1/2 falling into that category (**Figure 2A-B**). Remaining 64.2% of IGA3/4 patients are more difficult to distinguish from IGA1/2. There is a clear separation of the remaining IGA3/4 patients by the SCORAD Total Affected Area (**Figure 2C**).

**Figure 2.**
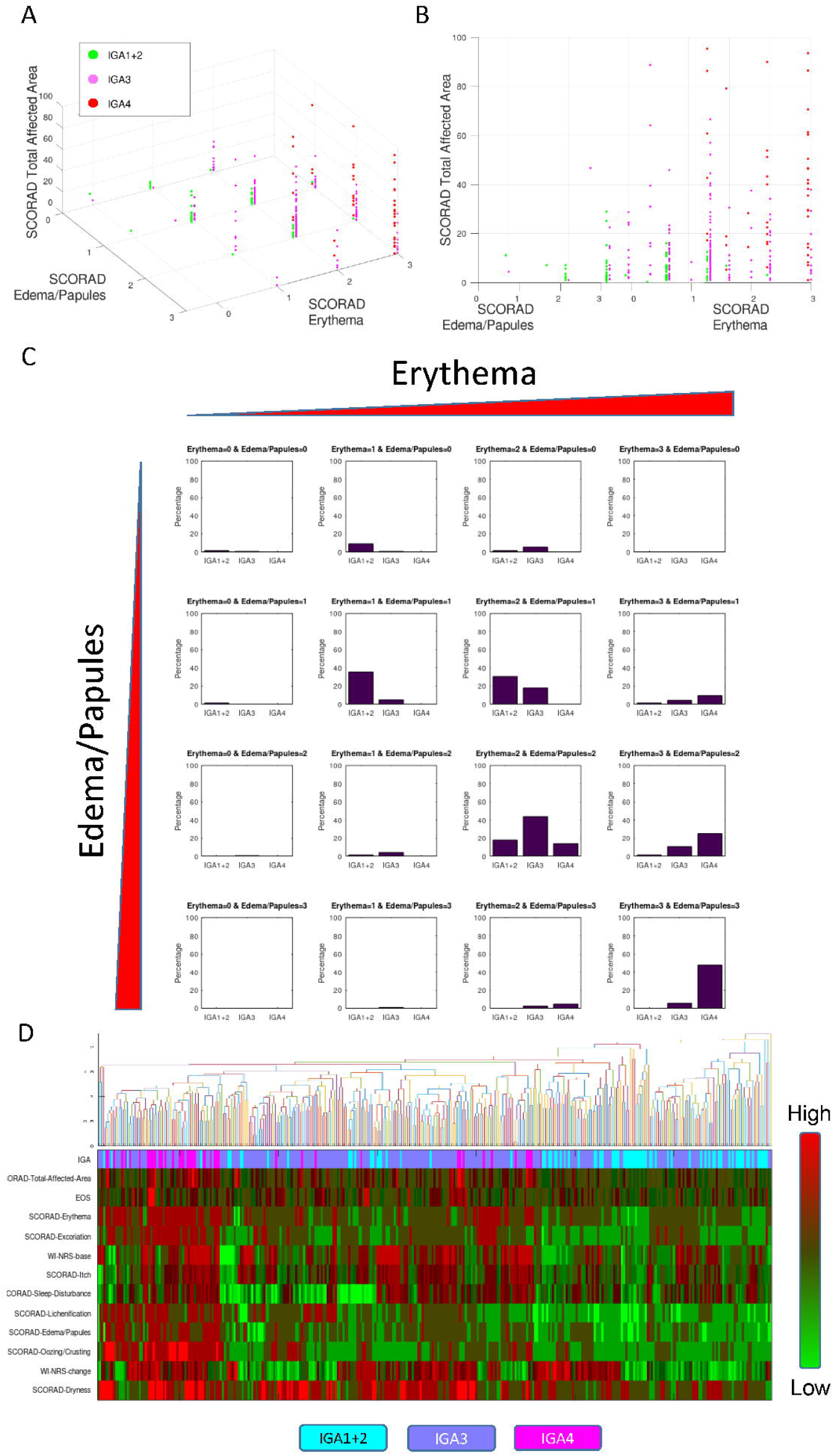
Differences between mild and moderate to severe subtypes in terms of clinical feature space.

We performed a series of classification and regression analyses to understand the underlying nature of the severity subgroups utilizing multiple clinical variables at once. We focus our analysis using classification trees. Based on a series of five clinical features from the SCORAD scale, we were able to accurately differentiate between IGA 1/2 versus IGA 3/4. We obtained a robust classification using SCORAD Total Affected Area (%), Edema/Papules, Erythema, Lichenification, and Excoriation. Five-fold cross-validation classification (J48) yields weighted 83.8% precision and 80.1% recall and a large AUC 0.830. **Supplementary Figure 1** displays the actual tree (J48) revealing the cutoffs and splits.

### Eosinophils and IgE

Motivated by the hypothesis that atopy is key in the expression of AD, we evaluated the role of counts of eosinophils and levels of IgE and their association with the clinical expression of AD. We evaluated lab values (EOS, IgE, systemic cytokines) that may differentiate milder from more severe AD. We detect a significant association between IGA and EOS levels. We have shown that more severe AD (IGA3/4) is associated with significantly higher counts of eosinophils as compared to milder AD (IGA1/2) (**Figure 1**), an effect that is significant (Mann-Whitney p<0.01). The effect is persistent in an extended screening cohort of 765 samples. Moreover, there is a significant difference between IGA 3 and 4 in the EOS levels (p<0.0008) and 1/2 vs. 4 (p<0.0001). This suggests that systemic eosinophil levels are increasingly correlated across the IGA severity scale. All but 3 cases of the 32 cases above 0.5 are moderate to severe AD subjects (29/282 of IGA3/4 and only 3/79 IGA1/2). Additionally, we report a significant difference in distribution of patients with elevated serum IgE (**Figure 1D**, ≥100 IU/mL, p<0.001).

### Genetic landscape of AD endotypes

We performed whole genome sequencing of the entire study population (n=765). We investigated the incidence of all *FLG* LOF variants in the genomes of the EPIONE study patients, and compared them with the whole genome sequences of a control population (n=316). of healthy volunteers as well as individuals from gnomAD database. More severe AD is associated with a higher prevalence of *FLG* LOF variants (p<0.0001, OR: 2.3, CI: 1.6-3.2). This is significant enrichment as compared to milder (p=0.99, OR: 0.89, CI: 0.3-2.0). Milder AD has incidence of LOF *FLG* variants comparable to normal population as defined by gnomAD and an internal set of controls (IGA1/2: 6.4%, Vanda Controls: 6.2%, GnomAD: 8.4%) whereas the moderate to severe subgroup has an elevated frequency of LOF *FLG* variants (IGA3/4: 17.7%). This is contrary to the general observation that AD cases have higher prevalence of *FLG* mutations than the control population, as this effect seems to be specific to IGA3/4 subjects. The variants detected are depicted on **Figure 3** which displays the location of individual LOF variants in both endotypes. We have now showed that the increased frequency can be ascribed to the severe AD subtypes. The results are consistent for EDC gene variants. Furthermore, we detect higher frequency if individuals with two or more FLG mutations in the severe group as well as incidence of cases with CNV within the *FLG* gene present in the IGA3/4. The allelic frequency of th**e** two prevalent *FLG* LOF variants, p.R501* and p.S761fs as well as the OR is displayed on panel B of **Figure 3**.

**Figure 3.**
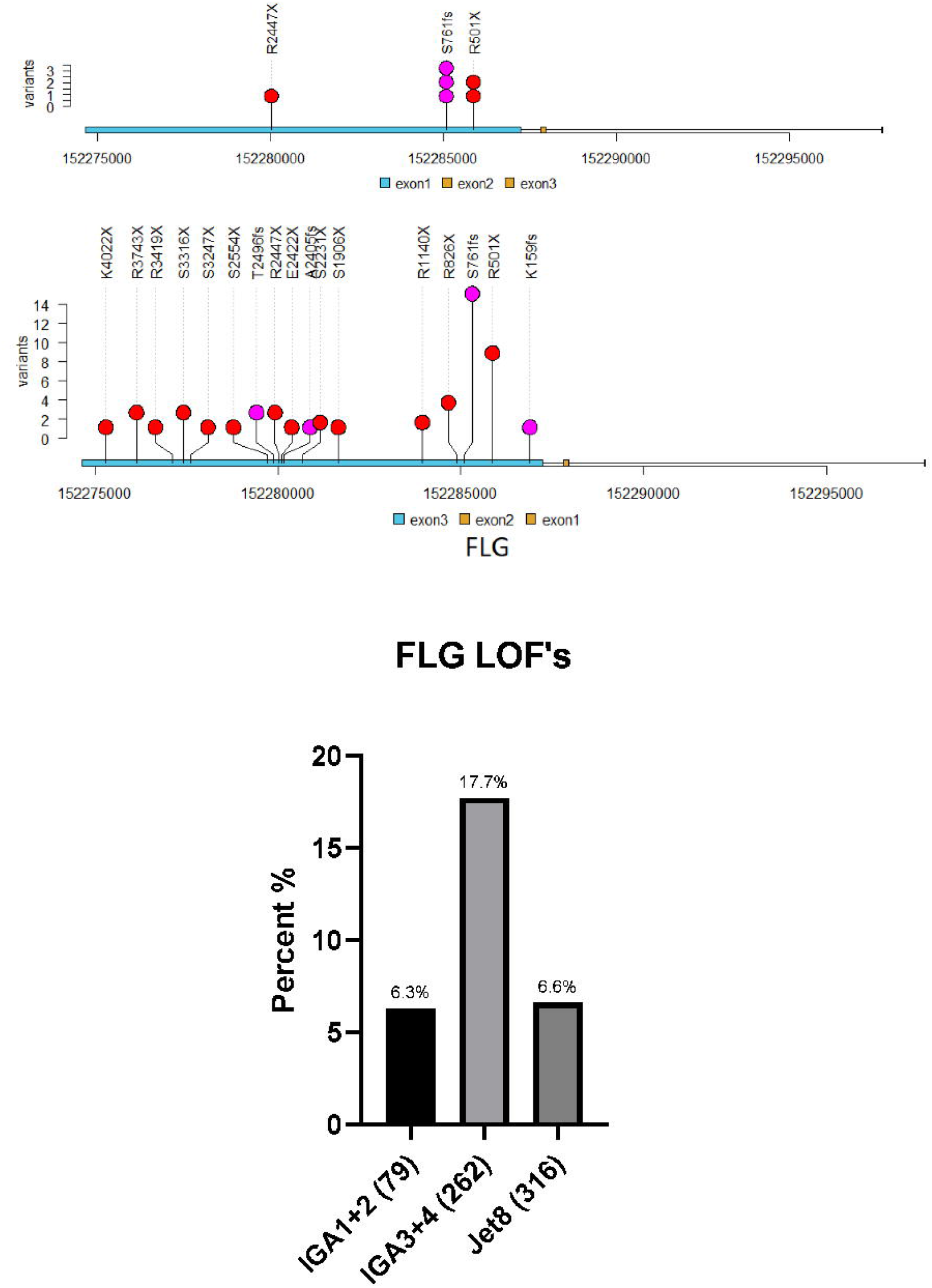
FLG LOF variants across IGA endotypes.

### *IL5RA* gene rare variants differentiate between IGA subgroups

We have performed *in silico* interleukin (IL) screen using gene-centric collapsing analysis on a pathway of IL genes as defined by GO terms, comprised of 96 genes. In a rare variant analysis we obtain a significant result after conservative multiple hypothesis testing in IL5RA as differentiating between the two severity groups. Direction of effect is consistent with a higher EOS levels observed in IGA3/4. Binding of IL-5 to the α-subunit of the IL5Ra promotes the heterodimerization of iL-5Rα and βc subunits. As a result, many signal transduction pathways are activated, including JAK/STAT modules, MAPK, Pi3K, and NF-κB. The stimulation of these kinases and TFs drives the expression of key genes responsible for differentiation, survival, degranulation, adhesion, and recruitment of eosinophils. Furthermore through whole genome sequence analysis we have also discovered that polymorphisms in the IL5Ra gene are associated with higher count of eosinophils in this population of AD patients. The IL5Ra rare variant rs200005614 associates with significantly higher level of EOS consistent with the effect observed. Previously other variants were found to be associated with EOS such as *IL5RA c*.*−5091G>A* which was shown to be associated with eosinophil count. *IL5RA* plays an important role in coordinating the release of eosinophil and IgE production against antigens leading to the development of atopy. The association of the IL5RA variants with the blood eosinophil counts and the severe IGA indicates that the IL5RA gene has a role for controlling crucial role in eosinophil responses.

### Cytokines

We measured a panel of systemic cytokines collected at baseline. We evaluated which cytokines at baseline are above the normal ranges (as defined by clinical standards^26^) and show the results on a heatmap of ratios to reference range. This is displayed on **Figure 4** (log2 ratio of cytokine to a maximum value reported in reference range, red being above threshold levels levels). The observed trends are suggestive of elevated levels of Th2 (IL-4, IL-5, and IL-13). Increased IL-5, as well as IL-4 and IL-23 form a distinct, predominant group of patients (**Figure 4**).

**Figure 4.**
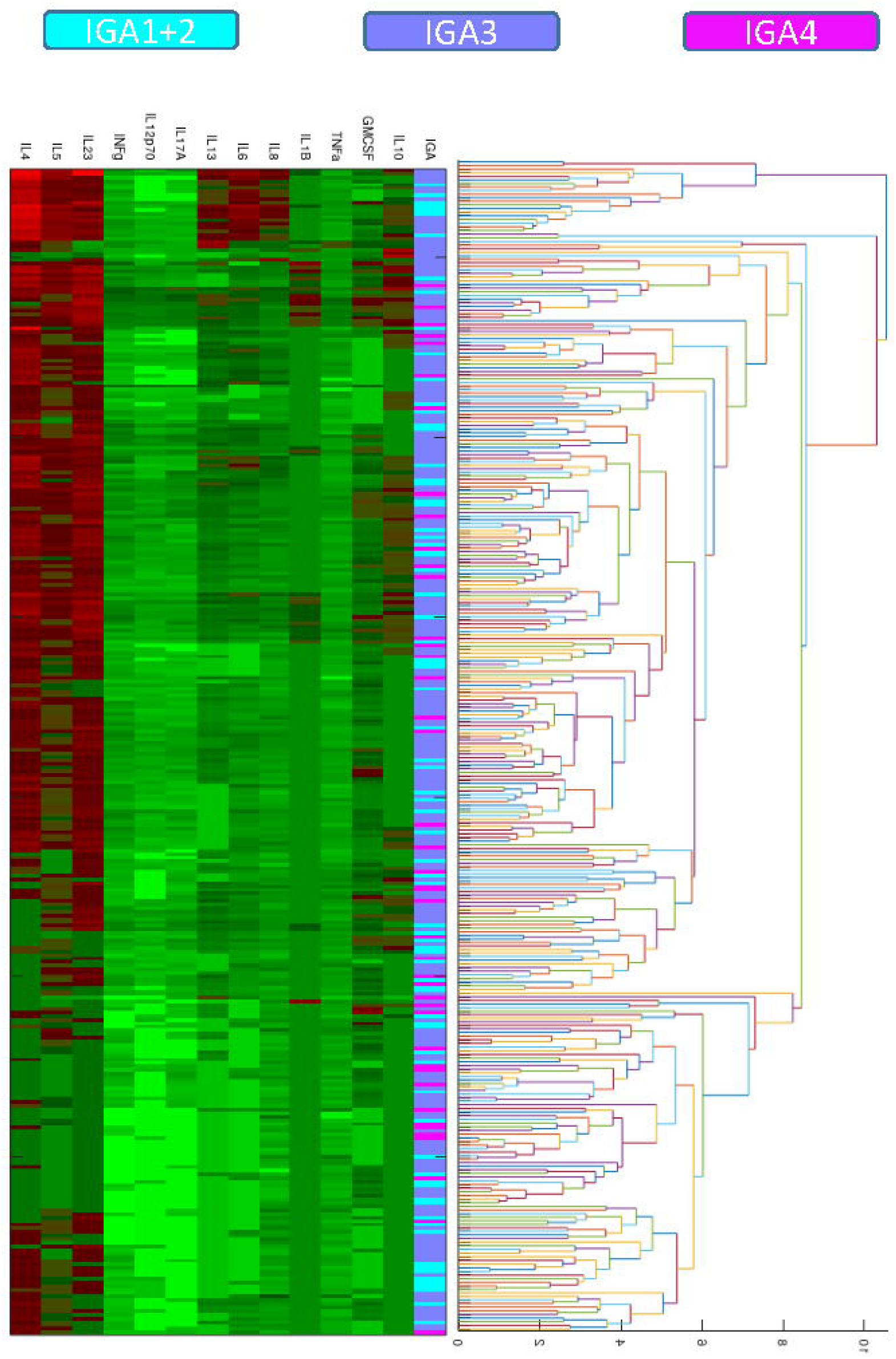
Cytokine panel at baseline (ratio to reference range)

Interestingly, there is a subgroup of patients that also have elevated IL-6, IL-8, and IL-13 (blue ribbon highlight on **Figure 4**). IL-5 is produced by type 2 T helper cells and mast cells. It stimulates B cell growth and increases immunoglobulin secretion, and primarily is the key mediator of eosinophil activation. Its high expression has been previously associated with chronic AD. It stimulates activated B cell and T cell proliferation, and differentiation of B cells into plasma cells. IL-4 has been implicated in tissue inflammation and epidermal barrier dysfunction^27^. These patients have decreased IFN-gamma and IL-12 levels across all patients, not limited to patients with elevated Th2. Whereas the main signature pattern (elevated IL-4, IL- 5, and IL-23) is still as abundant in IGAs 1/2 and even 3s, subjects defined as 4 lack a subgroup of patients with high level of IL-6, IL-8, and IL-13. **Figure 5** displays the cytokine levels in IGA subgroups. There is a distinct group of patients across all IGA severities that has low systemic levels of all cytokines. In addition, many more severe outliers have several magnitude higher levels across the entire cytokine panel as compared to milder cases.

**Figure 5.**
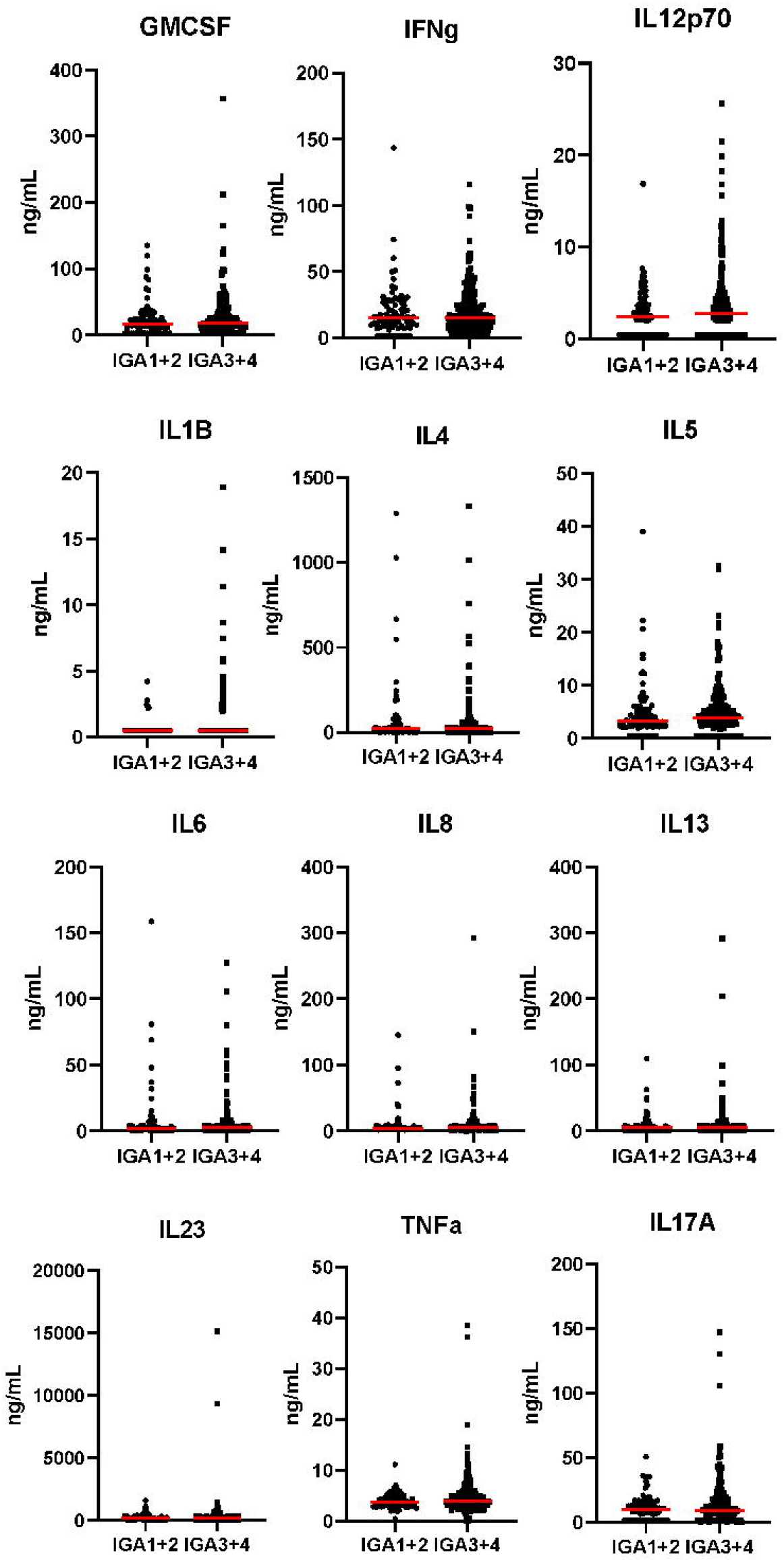
Individual cytokine scatter plots.

## Discussion

Defining AD subtypes has been a debated subject in the field. Definitions analogous to asthma have been previously used based on elevated IgE: with presence (“extrinsic”) or absence (“intrinsic”) of increased IgE^28^. Now, endotypes defined by different age groups, ethnicities and according to IgE levels and FLG mutation status are in the scope of discussions^8^.. Rare information is available regarding endotypes and different stages of AD in one population.

According to epidemiological studies, mild AD is the most common type of AD, affecting approximately 60% of AD casesThe factors underlying severity are not well understood. We present here clinical and genetic evidence supporting the concept of AD in different stages being composed of distinct endotypes. Clinical features suggest milder and severe AD appear to be distinct entities that can be differentiated by five clinical features from the SCORAD index. More severe AD is associated with a higher prevalence of mutation burden in the FLG skin barrier structural protein. These data are consistent with previous investigations, including a study in Denmark on (n=470) patients with AD^29^, as well as extensive studies of *FLG* gene^5^.

We found that more severe AD is associated with significantly higher count of eosinophils as compared to milder AD. Eosinophils are mediators of the inflammatory response and are responsible for recruiting other immune cells in the lesions. As such, it is likely that severe pruritus associated with both mild and severe AD may have different mediators (inflammation, skin barrier disturbance). Based on our findings we hypothesize that the contribution of SP in mild AD, in generating and transmitting the pruritic signal, may be relatively more significant than that seen in severe AD where inflammation dominates. Through whole genome sequence analysis we have also discovered that polymorphisms in the IL5Ra gene are associated with higher count of eosinophils in this population of AD patients. The association between eosinophils and IL5RA has been previously reported in asthmatic patients ^30^. Here we find the association by gene-centric testing between the two severity groups. This is in agreement with a reported association of rs2522411 variant in the IL-5 gene which was significantly associated with extrinsic AD^30^. When evaluating IL genes we detect IL5RA rare variants further differentiating between severities, with rare IL5RA variants accumulating in the more severe cases.

The described findings lend credence to the existence of endotypes within AD that are manifested clinically and are becoming more understood in terms of underlying mechanisms. These findings may have implications for both diagnosis and treatment course of the disorder. As seen in (Welsh et al. 2020, unpublished) a robust anti-pruritic effect was observed in only the IGA1/2 population while the difference from placebo was not significant in the more severe IGA population. Endotype analyses define the underlying clinical and genetic factors of mild versus more severe AD, and would indicate that the immune response and skin barrier dysfunction tends to be increased in more severe AD. This may mean treating such patients may require more immunosuppressive/immunemodulating therapies. In contrast, mild patients without such underlying factors may be better served with direct targeted therapies such as an anti-pruritic agent addressing the severe itch they may still experience despite their lesions being almost clear or mild. Further attempts to define AD endotypes should be made to define the optimal therapeutic approach, moving towards precision medicine-based treatment of AD based on clinical, molecular, and genetic characteristics.

## Data Availability

Data available upon request

## Acknowledgements

We are grateful to all the study participants and all whom made the study possible

## Abbreviations

AD: Atopic Dermatitis
EDC: Epidermal Differentiation Complex
EOS: Eosinophils
FLG: Filaggrin
IGA: Investigator’s Global Assessment
IL: Interleukin
LOF: Loss-of-Function
NK1R: Neurokinin-1 Receptor
CORAD: SCORing Atopic Dermatitis
SP: Substance P
vIGA-AD: Validated Investigator Global Assessment for Atopic Dermatitis

